# Post-lockdown detection of SARS-CoV-2 RNA in the wastewater of Montpellier, France

**DOI:** 10.1101/2020.07.08.20148882

**Authors:** Julie Trottier, Regis Darques, Nassim Ait Mouheb, Emma Partiot, William Bakhache, Maika S. Deffieu, Raphael Gaudin

## Abstract

The evolution of the COVID-19 pandemic can be monitored through the detection of SARS-CoV-2 RNA in sewage. Here, we measured the amount of SARS-CoV-2 RNA at the inflow point of the main waste water treatment plant (WWTP) of Montpellier, France. We collected samples 4 days before the end of lockdown and up to 45 days post-lockdown. We detected increased amounts of SARS-CoV-2 RNA at the WWTP, which was not correlated with the number of newly diagnosed patients. Future epidemiologic investigations may explain such asynchronous finding.

## Main text

SARS-CoV-2 is the etiologic agent responsible for the current coronavirus disease 2019 (COVID-19) pandemic. Wastewater-based epidemiology represents an attractive strategy to surveil the evolution of virus circulation in populations (Bivins et al., 2020, Carducci et al., 2020), contributing to cost-effective virus control without infringing on individual liberties. About half of symptomatic patients shed SARS-CoV-2 RNA in their stools, with persistent fecal viral shedding reported (Chen et al., 2020, Wang et al., 2020, Wu et al., 2020, Xiao et al., 2020, Xu et al., 2020). Recently, an asymptomatic child was reported as negative for SARS-CoV-2 RNA based on throat swab specimen, while his stools were positive (Tang et al., 2020), suggesting that symptomatic and asymptomatic persons are likely to release SARS-CoV-2 RNA in city sewerages. Indeed, several reports indicate that SARS-CoV-2 RNA was readily detected in wastewater, and it is proposed that such approach could anticipate the occurrence of novel COVID-19 outbreaks in low prevalence regions (Ahmed et al., 2020, La Rosa et al., 2020, Medema et al., 2020, Orive et al., 2020, Randazzo et al., 2020). The end of the stringent lockdown (that occurred in France on May 11^th^) is therefore an adequate time to measure the re-emergence of the virus through the monitoring of wastewater.

Here, we collected effluent composite samples (using a 24-hour automatic sampler) in waste water upstream of the main waste water treatment plant (WWTP) of Montpellier metropolitan area located in Lattes, France. The sampling dates were May 7^th^, 18^th^, 26^th^, June 4^th^, 15^th^ and 25^th^, to monitor SARS-CoV-2 RNA expression levels during lockdown and up to 45 days after its end. At this time, the virus was still circulating in the area, but the incidence was low (number of newly diagnosed COVID-19 patients per day < 20).

Collected wastewater was processed as follows: on the day of water collection, samples were maintained at 4°C for transport and immediately cleared by centrifugation at 4500 g for 30 min at 4°C. The supernatant was passed through a 40 µm cell strainer (Corning) to remove large floating components. At this stage, the samples were frozen at −20°C for later analyses with samples collected at other timepoints. Upon thawing, RNAs were concentrated on a Vivaspin 50 kDa MWCO filter membrane (Sartorius). Starting from 50 ml of water, the sample was concentrated down a hundred times to an adjusted volume of 500 µl. RNA extraction using the NucleoSpin RNA Virus kit (Macherey-Nagel) and RT-qPCR was performed on 10 µl of purified RNA using the TaqPath One-Step RT-qPCR, CG master mix (ThermoFisher Scientific). The N1 and N3 primer/probe sets designed by the center for disease control (CDC) were used to detect SARS-CoV-2 RNA and a standard curve was run in parallel using a positive control plasmid (Integrated DNA Technologies) coding for the nucleoprotein (N) of SARS-CoV-2. Using RNA extracted from Vero E6 cells either non-infected or infected with SARS-CoV-2 *in vitro*, we showed that the N1 and N3 primer/probe sets recognized solely the RNA from infected cells (Table 1). A recent study used a Dengue virus (DENV) sequence surrogate to determine PCR efficiency (Medema et al., 2020) but in Montpellier area, this approach would be risky, as autochthonous DENV infections have been repeatedly reported in the south of France (European Centre for Disease Prevention and Control). In order to determine the RT-qPCR efficiency intrinsic to each sample, we used a sensitive primer/probe set previously described (Ro et al., 2017) that target the VP40-encoding RNA of Ebola virus (Zaire strain). First, we showed that the Ebola standard (Ebo Std) primer/probe set was not detecting RNA from SARS-CoV-2-infected Vero E6 cells (Table 1). Using water samples from upstream WWTP of the Montpellier metropolitan area collected on June 15^th^, we found that the Ebo Std primer/probe set gave no signal while the primer/probes N1, N3, and RLP27 (targeting the human *rlp27* gene) returned positive signals (Table 1).

**Table 1.** Specificity of the primer/probe sets from *in vitro* SARS-CoV-2 infected cells. The Table shows the cycle threshold (Ct) of individual RT-qPCR reaction. “No Ct” indicates that no signal was detected over 40 cycles.

| Primer/probe set | N1 | N3 | Ebo Std | RLP27 |
| --- | --- | --- | --- | --- |
| RNA from non-infected cells | No Ct | No Ct | No Ct | - |
| RNA from SARS-CoV-2 infected cells | 19 | 17.4 | No Ct | - |
| RNA from waters collected upstream of the WWTP | 36.4 | 37.6 | No Ct | 30.9 |
|  | No Ct | 37.3 | No Ct | 30.2 |

A synthetic RNA sequence of 91 nucleotides (gcagaucgaauccacucaggccaaucuauggugaaugucauaucgggccccaaagugcuaaugaaguuuggcuuccucuaggugucggcag) derived from the VP40-coding gene of Ebola virus and recognized by the above-mentioned Ebo Std primer/probe set was purchased (Integrated DNA Technologies) to be used as an internal normalization standard. Standard curves were run using the N control plasmid (N1 and N3 primer/probe sets) or the Ebo Std RNA (Ebo Std primer/probe set) to estimate copy numbers (Figure 1). Of note, the Ebo Std primer/probe set was less sensitive than N1 and N3, probably because the Ebo Std synthetic template RNA is relatively fragile and short (91 nucleotides).

**Figure 1.**
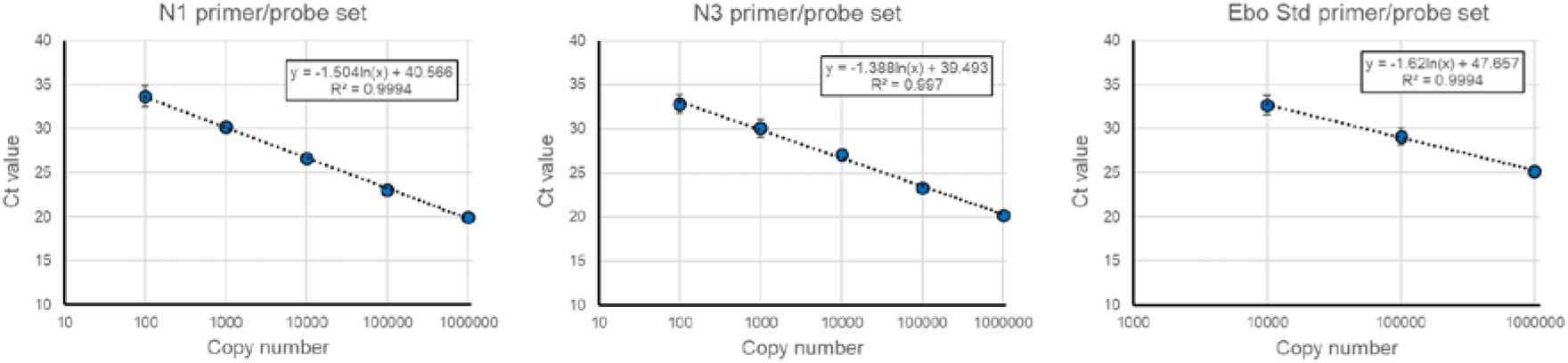
Standard curves of the N1 (left), N3 (middle) and Ebo Std (right) primer/probe sets showing the cycle threshold (Ct) value at indicated template copy number.

Next, we evaluated the relative efficiency of RNA extraction and RT-qPCR reaction for each sample by adding 10^7^ copies of synthetic Ebo Std RNA before RNA extraction. This step allows quantitative comparison of samples collected at different dates. Our data show that RNA extraction and RT-qPCR (RERP) efficiency was marginally affected according to the collection date (Table 2).

**Table 2.**
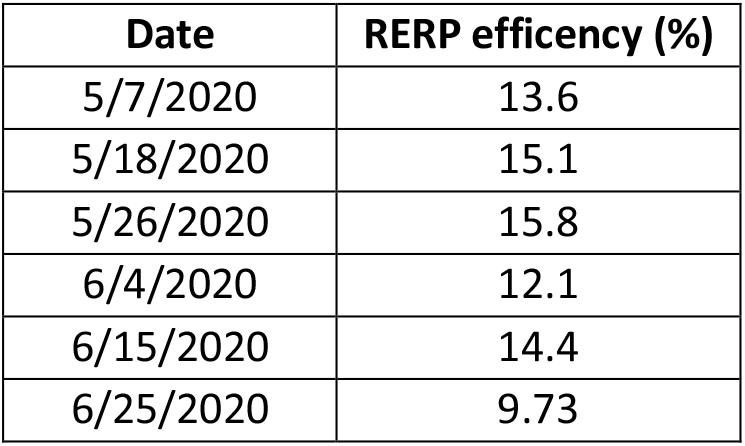
RNA extraction and RT-qPCR (RERP) efficiency using Ebo Std. RERP efficiency was calculated by dividing the copy number of Ebo Std RNA measured by RT-qPCR (using the standard curve in Figure 1) by the theoretical copy number of synthetic Ebo Std RNA and multiplied by 100.

| Date | RERP efficiency (%) |
| --- | --- |
| 5/7/2020 | 13.6 |
| 5/18/2020 | 15.1 |
| 5/26/2020 | 15.8 |
| 6/4/2020 | 12.1 |
| 6/15/2020 | 14.4 |
| 6/25/2020 | 9.73 |

Next, we measured the SARS-CoV-2 RNA levels using N1 and N3 primer/probe sets in wastewater collected upstream of the main WWTP of the Montpellier metropolitan area on May 7^th^, 18^th^, 26^th^, June 4^th^, 15^th^ and 25^th^ (Figure 2A). Of note, the Montpellier wastewater network is partially united and thus, we checked that rain precipitation was negligible and would not significantly dilute the samples according to the collection date (Figure 2B).

**Figure 2.**
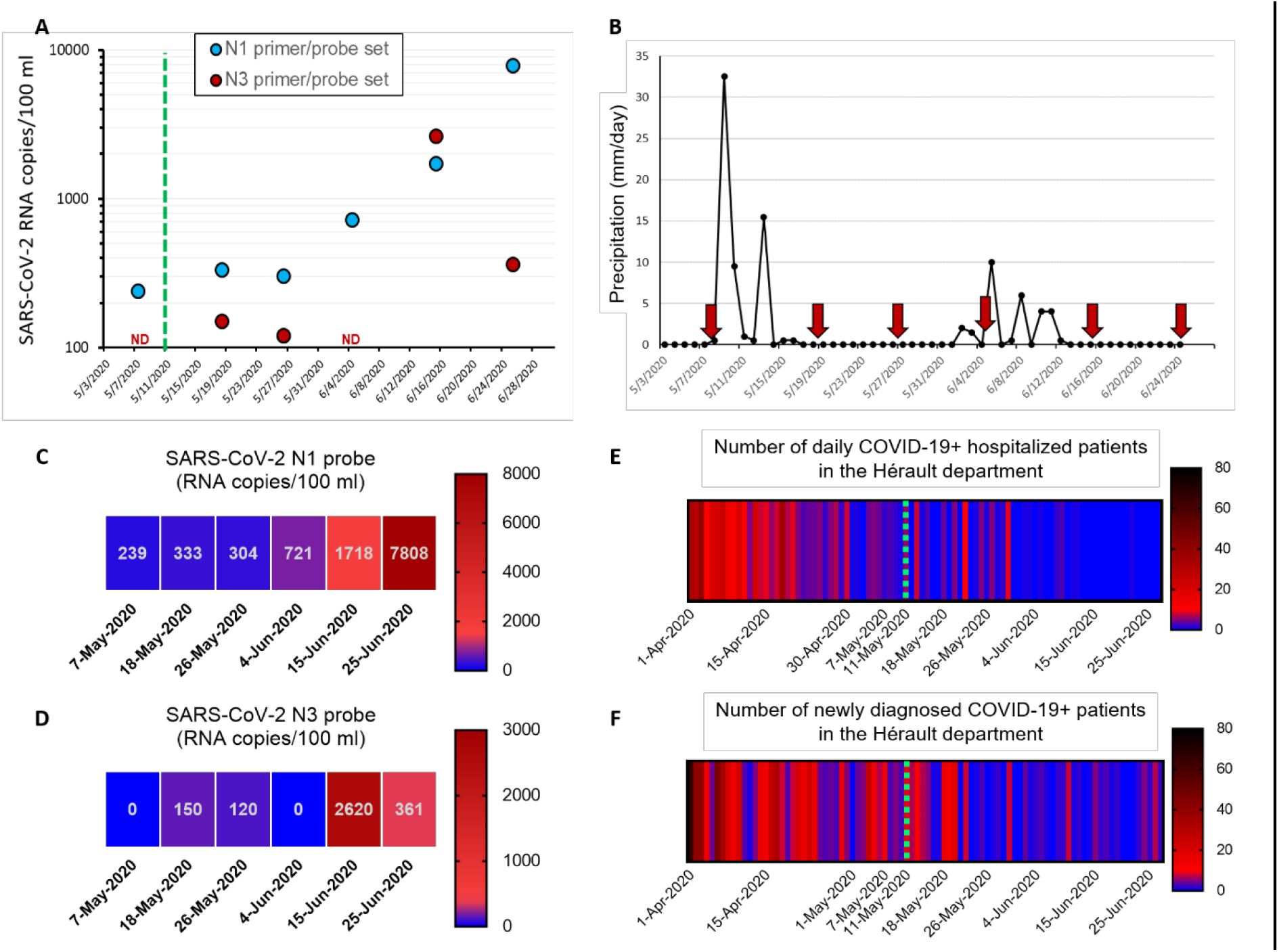
SARS-CoV-2 RNA detection in the Montpellier wastewater and number of COVID-19 cases. (A) The number of SARS-CoV-2 RNA copies was measured using either the N1 or N3 primer/probe set. Each dot corresponds to the concentration of SARS-CoV-2 RNA in wastewater measured from the sum of the RNA copy number calculated from two RNA extractions and four RT-qPCR reactions performed in two individual runs. Sterile water wells were included in duplicates for each primer/probe set in each plate as negative control and always returned “No Ct” (not shown on the graph). Of note, the graph does not take into account the RERP efficiency calculated in Table 2. The green dotted line indicates the end of the strict lockdown in France. ND: non-detected. (B) The graph shows precipitation in millimeter per day recorded in Montpellier (station Lavalette 34172005). The red arrows indicate our wastewater collection dates. (C-D) Heatmaps of the SARS-CoV-2 RNA concentration calculated in A specific to the N1 (C) and N3 (D) primer/probe sets. (E-F) The heatmaps show the number of daily COVID-19 patient hospitalized (E) and the number of newly confirmed COVID-19 cases (F) in the Hérault department (source: Santé Publique France). The green dotted line indicates the end of the strict lockdown in France.

Our data highlights that the wastewater from the two later dates (June 15^th^ and 25^th^) contained more viral RNA than the ones from previous dates (Figure 2A and color-coded in Figure 2C-D). In contrast, the number of daily COVID-19 patients hospitalized in the Hérault department (> 40% inhabitant living in the Montpellier metropolitan area) and the newly diagnosed patients (Figure 2E-F) is overall decreasing since April 1^st^. From these observations, it seems that there is no direct temporal correlation between SARS-CoV-2 RNA detection in wastewater and the epidemiological features associated with COVID-19, at least on such a short time window. This intriguing result is reminiscent of a recent Spanish study, in which the authors could detect SARS-CoV-2 RNA in wastewater weeks before the first COVID-19 cases were reported (Randazzo et al., 2020). However, they could not see a correlation between SARS-CoV-2 RNA levels in wastewater and the number of newly diagnosed COVID-19 patients. On the same line, Medema & colleagues showed a correlation between the cumulative cases of COVID-19 and SARS-CoV-2 RNA although the data were not correlated as a function of time (Medema et al., 2020).

In conclusion, we report effective detection of SARS-CoV-2 RNA in the wastewater of Montpellier area upstream of the treatment plant and identified a recent increase of the amount of detected viral RNA. At this stage, we are unable to determine whether this increase is due to an upcoming increase of COVID-19 cases in the area or to intrinsic SARS-CoV-2 RNA variations associated with uneven virus shedding (from patient-to-patient and depending on the stage of the disease for a given patient). Moreover, various other parameters might also impact these results, such as people from distant clusters moving to second homes and tourist accommodation, the chronic underestimation of prevalence rates, or local variability in the geographical pattern of virus spread. Both hypotheses are non-exclusive and future multiparametric investigations are required to better determine whether the monitoring of wastewater could be a powerful predictive tool to future epidemic outbreaks.

## Data Availability

Data are available upon request

## Authors’ contributions

J.T established our consortium and initiated the project. J.T and R.G designed the study. R.D provided epidemiological datasets and geomatics expertise. N.A.M provided the rain precipitation dataset and hydrology expertise. E.P, W.B, M.D and R.G performed and optimized the technical experimentations. R.G wrote the manuscript and J.T, R.D, N.A.M and W.B edited and commented on the manuscript. All authors agree on the final version of the manuscript.

## Acknowledgements

We thank Jean Verdier and Nima Machouf for their helpful comments and suggestions, and Yonis Bare for technical assistance. The strain Beta CoV/France/IDF0372/2020 was supplied by the National Reference Centre for Respiratory Viruses hosted by Institut Pasteur (Paris, France) and headed by Dr. Sylvie van der Werf.

## Conflict of interest

None declared

## Funding statement

This work was funded by the CNRS. The funder had no role in the design of the study.

## Notes

### Competing Interest Statement

The authors have declared no competing interest.

### Author Declarations

All relevant ethical guidelines have been followed

